# Racial and Socioeconomic Disparities in Blood Pressure Control Before and After Intracerebral Hemorrhage

**DOI:** 10.64898/2026.05.11.26352899

**Authors:** Samuel Namian, Joel Smith, Sofia Constantinescu, Yome Tawaldermedhen, Santiago Clocchiatti-Tuozzo, Cyprien A. Rivier, Shufan Huo, Kane Wu, Victor Torres Lopez, Sanjula Dhillon Singh, Christopher D. Anderson, Jonathan Rosand, Seyedmehdi Payabvash, Santosh Murthy, Kevin N. Sheth, Adam de Havenon, Guido J. Falcone

## Abstract

**Background:** Hypertension is the most potent modifiable risk factor for recurrent intracerebral hemorrhage (ICH), yet blood pressure (BP) control after ICH remains suboptimal, particularly among disadvantaged racial and socioeconomic groups. To what extent post-ICH BP disparities reflect pre-existing hypertension inequities versus differences in post-ICH management is unknown. We examined disparities in BP control before and after ICH, assessed whether post-ICH care differentially improves BP across groups and whether post-ICH disparities persist after accounting for pre-existing BP differences.

**Methods:** We performed a case-only study in the All of Us Research Program, identifying ICH survivors using electronic health record diagnosis codes. Mean systolic BP was calculated for pre-ICH (1–365 days before) and post-ICH (30–365 days after) windows. Neighborhood deprivation tertiles were calculated using 3-digit ZIP codes. The primary outcome was uncontrolled BP (≥140 mmHg). Logistic regression estimated odds of uncontrolled BP, and mediation analysis estimated the proportion of post-ICH disparities explained by pre-ICH BP.

**Results:** Among 2,226 ICH survivors (mean age 60; 50.6% female), 1,760 had pre-ICH and 1,852 had post-ICH BP data. Uncontrolled BP was more common in Black than White survivors both pre-ICH (38.9% vs 21.4%; p<0.001) and post-ICH (34.3% vs 16.3%; p<0.001), and in Deprived versus Privileged neighborhoods post-ICH (23.7% vs 15.8%; p<0.001). In adjusted models, Black race (OR 3.51; 95% CI 2.55–4.83; p<0.001) and Deprived neighborhoods (OR 1.38; 95% CI 1.00–1.91; p=0.048) were associated with uncontrolled post-ICH BP. Among survivors uncontrolled before ICH, 67% of White but only 45% of Black survivors achieved control afterward (p=0.001). Adjusting for pre-ICH BP control status only modestly attenuated the Black-White disparity (OR 4.05 to 2.95; P<0.001). In mediation analyses, pre-ICH BP explained only 27% of the racial (P<0.001) and 26% of the deprivation (P=0.014) disparity.

**Conclusions:** Racial and socioeconomic disparities in BP control persist after ICH, but most post-ICH disparities are not explained by pre-existing inequalities. More advantaged populations achieve greater BP improvement, suggesting effective post-ICH management exists but does not reach all patients equitably. Targeted interventions addressing barriers to post-ICH BP control in disadvantaged populations may substantially reduce persistent disparities.

## Background

Intracerebral hemorrhage (ICH) is the deadliest form of stroke and a leading cause of neurological disability. Hypertension is the leading modifiable risk factor for both primary and recurrent spontaneous, non-traumatic ICH, and intensive blood pressure (BP) lowering after stroke reduces recurrent vascular events.^1–3^ Despite widespread recognition that BP control is the dominant modifiable determinant of ICH recurrence and that it is achievable with effective antihypertensive medications, fewer than half of ICH survivors reach recommended BP targets at 3-months post-ICH.^3–6^ Understanding the mechanisms that drive poor BP control and identifying gaps in hypertension management after ICH is essential to develop targeted interventions.

Hypertension disparities between racial and socioeconomic groups are well established. Black adults have the highest hypertension prevalence of any racial or ethnic group in the United States, and despite similar rates of antihypertensive medication use, remain substantially less likely to achieve BP control than White adults.^7–11^ Residents of socioeconomically disadvantaged neighborhoods also face higher hypertension incidence and poorer BP control,^12,13^ and in the REGARDS study, social determinants of health mediated approximately one-third of the Black-White disparity in uncontrolled BP among treated adults.^14^ These population-level disparities extend to stroke survivors: Black stroke survivors have significantly lower rates of BP control despite similar medication use,^15^ and evidence suggests that minority survivors receive less guideline-concordant secondary prevention and face greater barriers to physician access and medication affordability.^16,17^ Racial differences in antihypertensive prescribing patterns have been documented among stroke survivors specifically,^18^ and a standardized post-stroke management protocol reduced some racial differences in vascular risk factor control,^19,20^ suggesting that post-stroke disparities may not simply reflect pre-existing inequities but may also be shaped by differences in post-stroke care delivery.

In the context of ICH specifically, there are substantial racial disparities in BP management, recurrence, and mortality. ICH incidence among Black adults is approximately double that of White adults,^21^ and Black ICH survivors have significantly higher rates of recurrent hemorrhage, with differences partly explained by post-ICH BP control disparities.^22,23^ In a single-center cohort of 336 ICH patients, only 28% of survivors achieved substantial BP reduction in the year following ICH, and minority survivors and those from disadvantaged neighborhoods were significantly less likely to improve.^24^ However, pre-stroke hypertension status also strongly predicted post-stroke BP control even after accounting for race and socioeconomic status, raising the question of whether post-ICH BP disparities primarily reflect pre-existing inequities or also reflect differences in post-ICH management.^24^ That predominantly White, single-center study lacked the sample size and diversity to formally decompose these contributions.

Whether pre-existing disparities or differences in post-ICH management drive the gap has direct clinical implications. If pre-existing disparities account for most of the post-ICH inequality, then reducing post-ICH BP disparities may require first addressing broader population-level hypertension inequities. However, if advantaged populations achieve meaningful BP improvement after ICH while disadvantaged populations do not, that would suggest effective post-ICH management exists but is not reaching all patients equitably, and that targeted post-stroke interventions could meaningfully close the gap. To address this question, we leveraged the NIH All of Us Research Program,^25^ which provides longitudinal electronic health record-derived BP measurements before and after ICH within the same individuals across a large, diverse U.S. cohort. We aimed to characterize racial and socioeconomic disparities in BP control before and after ICH, examine whether post-ICH care differentially improves BP control across racial and socioeconomic groups, and quantify the extent to which pre-ICH BP mediates post-ICH hypertension control disparities.

## Methods

### Data Availability

Data are available from the All of Us Research Program for researchers with controlled tier access.

### IRB/Ethics Statement

We conducted a retrospective case-only study using data from the All of Us Research Program. We used the latest available data release (v8, Controlled Tier C2024Q3R4; Registered Tier R2024Q3R3), for which data collection was performed between May 2017 and October 2023. The study protocol was approved by the All of Us institutional review board, and participants or their legal guardians provided written informed consent.

### Study Population

The All of Us Research Program is a longitudinal cohort study that aims to enroll one million or more participants from across the United States, with emphasis on recruiting individuals from communities historically underrepresented in biomedical research.^25^ Participants contribute electronic health record (EHR) data, self-reported survey responses, physical measurements, and biospecimens.

To build our dataset, we first queried all systolic blood pressure measurements available in All of Us and excluded values less than 70 mmHg or at least 300 mmHg as likely erroneous. After these exclusions, 477,021 participants had at least one plausible systolic BP value from the total 633,547 enrolled participants. From this pool, we identified 3,265 participants with spontaneous intracranial hemorrhage using ICD-10 diagnosis codes for non-traumatic intracerebral hemorrhage (I61.), nontraumatic subarachnoid hemorrhage (I60.), and nontraumatic subdural hemorrhage (I62.0).

We then applied the following inclusion criteria: age 18 years or older at ICH; survival of at least one year after ICH, and at least one valid systolic BP measurement in either the pre-ICH window (1-365 days before ICH) or post-ICH window (30-365 days after ICH). After these restrictions, 2,226 ICH survivors met inclusion criteria. Of these, 1,760 (79%) had pre-ICH BP data, 1,852 (83%) had post-ICH BP data, and 1,386 (62%) had data in both windows. The study population selection process is summarized in Figure 1. 93 participants (4.2%) were missing neighborhood deprivation data, but all participants had age, sex, and race/ethnicity data available. Adjusted regression models required complete covariate data, and those that included deprivation index as an exposure or covariate excluded participants missing deprivation index (67 pre-ICH; 69 post-ICH). Change-in-status, differential BP improvement, and mediation analyses were further restricted to participants with data in both windows and by exposure group and pre-ICH BP status as described below.

**Figure 1.**
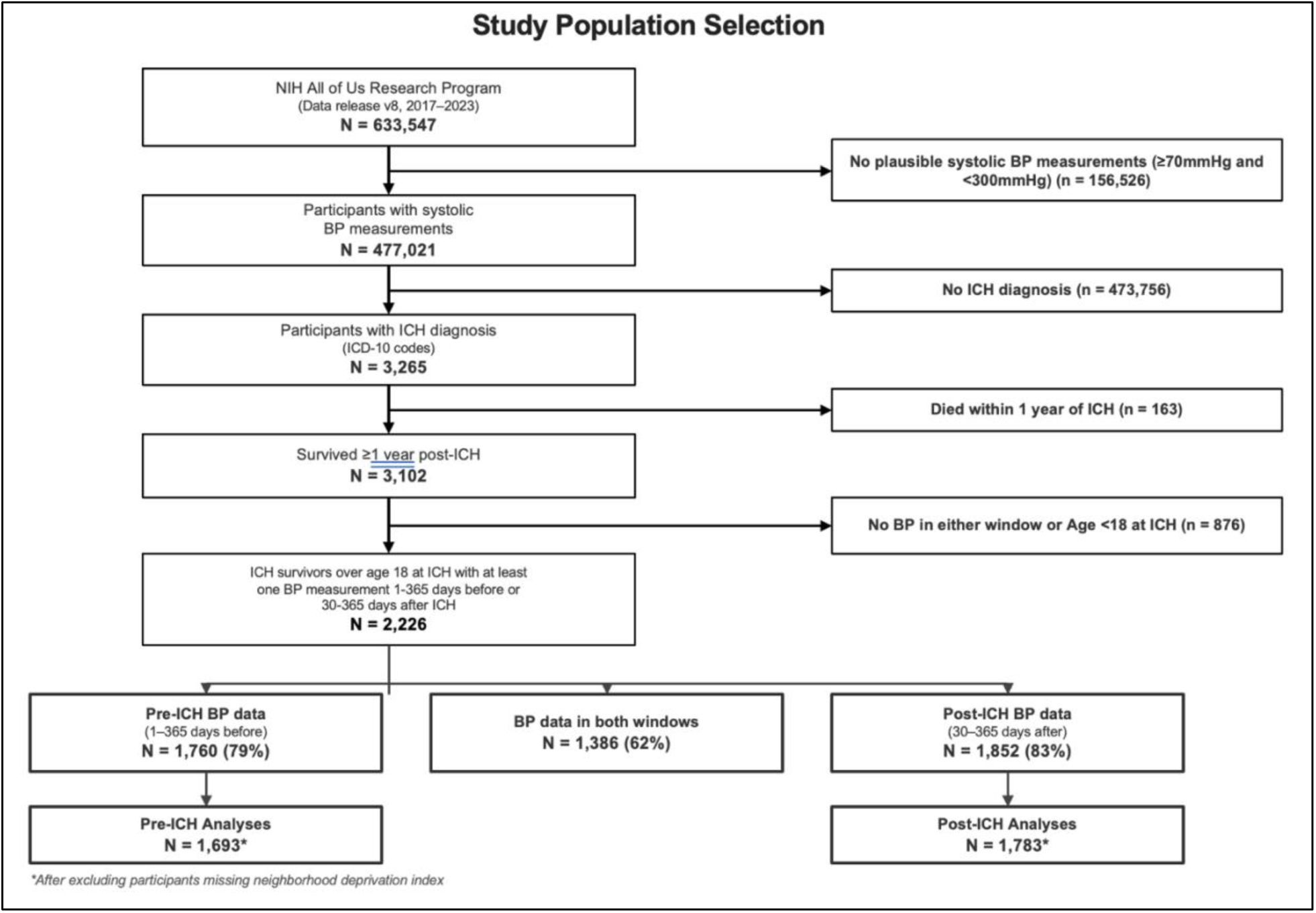
Study population selection. Flow diagram showing identification and inclusion of intracerebral hemorrhage (ICH) survivors from the NIH All of Us Research Program (data release v8, 2017–2023). From 633,547 enrolled participants, 477,021 had at least one plausible systolic blood pressure (BP) measurement (≥70 and <300 mmHg). From this pool, 3,265 participants with ICH diagnosis codes were identified. After excluding participants who died within one year of ICH (n=163), were under age 18 at ICH, or lacked BP data in both analytic windows, 2,226 ICH survivors met inclusion criteria. Of these, 1,760 (79%) had pre-ICH BP data (1–365 days before ICH), 1,852 (83%) had post-ICH BP data (30–365 days after ICH), and 1,386 (62%) had data in both windows. Adjusted regression analyses excluded participants missing neighborhood deprivation index data, yielding analytic samples of 1,693 for pre-ICH and 1,783 for post-ICH models. Change-in-status, differential BP improvement, and mediation analyses were restricted to participants with data in both windows. *Asterisk indicates sample sizes after excluding participants missing neighborhood deprivation index.

### Exposures

Exposures included self-reported race/ethnicity and neighborhood deprivation. Race/ethnicity categories were defined as non-Hispanic White, Black, Hispanic/Latino, and Other. These categories were mutually exclusive. Black participants that identified as Hispanic/Latino were categorized as Black, and White or Other participants that identified as Hispanic/Latino were categorized as Hispanic/Latino. Participants that did not identify as one of those three racial or ethnic groups were grouped into an “Other” category due to small sample sizes. Neighborhood deprivation was measured using the Nationwide Community Deprivation Index,^26^ which uses principal components analysis of six census-based socioeconomic indicators (fraction of the population below the poverty level, with less than a high school education, without health insurance, unemployed, receiving supplemental nutrition assistance, and with crowded housing) linked to participants via 3-digit ZIP codes. We categorized the index into tertiles (Privileged, Intermediate, Deprived) relative to our cohort.

### Covariates

Age at ICH was calculated from participant date of birth and the date of first spontaneous ICH diagnosis code. Sex was obtained from survey responses, and each participant was categorized as Male or Female/Other. To characterize the study population, we ascertained comorbidities including hypertension, diabetes mellitus, atrial fibrillation, chronic kidney disease, heart failure, chronic obstructive pulmonary disease, and prior cardiovascular events (ischemic stroke, myocardial infarction) using ICD-9, ICD-10, and SNOMED codes from the EHR. Annual household income, education level, insurance type, and smoking status were obtained from self-reported survey data. These variables were used to describe the cohort and were not included as covariates in regression models, which adjusted for age, sex, and the alternate exposure (neighborhood deprivation in race models; race/ethnicity in deprivation models) as described below.

### BP Measurement and Outcomes

For each analytic window (1-365 days before ICH and 30-365 days after ICH), we calculated each participant’s mean systolic BP from all available measurements. The 30-day post-ICH exclusion window was used to avoid capturing acute inpatient BP management, which may not reflect outpatient hypertension control. The primary outcome was uncontrolled BP, defined as mean systolic BP of at least 140 mmHg,^27,28^ irrespective of treatment status. Sensitivity analyses evaluated thresholds of 120, 130, and 150 mmHg.

### Statistical Analysis

All analyses were conducted using R version 4.3.^29^

**Pre-ICH and post-ICH BP control.** Unadjusted proportions of uncontrolled BP were compared between groups using pairwise chi-square tests (Black vs White, Hispanic/Latino vs White, and Other vs White for race; Intermediate vs Privileged and Deprived vs Privileged for deprivation). Multivariable logistic regression models estimated odds ratios (ORs) and 95% confidence intervals (CIs) for the association between each exposure and uncontrolled BP. For both race and deprivation models, we first adjusted for age at ICH and sex, and then additionally adjusted for neighborhood deprivation (in race models) or race/ethnicity (in deprivation models) to assess how each association changed after accounting for the other exposure. Covariates were selected based on established confounding relationships rather than statistical selection procedures. To assess whether post-ICH disparities persist after accounting for pre-existing BP differences, we repeated post-ICH logistic regression models with pre-ICH BP control status (uncontrolled vs controlled) as an additional covariate, restricted to the 1,386 survivors with BP data in both windows. To illustrate the combined burden of race and neighborhood deprivation, we created a joint four-level exposure variable (White-Privileged, White-Deprived, Black-Privileged, Black-Deprived) and estimated odds of uncontrolled BP with White-Privileged as the reference, adjusted for age and sex.

**Sensitivity analyses.** Adjusted logistic regression models were repeated at thresholds of 120, 130, and 150 mmHg to assess the robustness of findings across definitions of uncontrolled BP. As a sensitivity analysis, we repeated primary adjusted regression and mediation analyses restricting the cohort to intraparenchymal hemorrhage only.

**Change in BP control status.** Among survivors with BP data in both analytic windows, we stratified participants by pre-ICH BP control status (controlled: mean SBP <140 mmHg; uncontrolled: mean SBP >=140 mmHg) and examined transitions in BP control after ICH. For survivors with uncontrolled pre-ICH BP, we modeled odds of achieving BP control after ICH by exposure. For survivors with controlled pre-ICH BP, we modeled odds of losing BP control after ICH by exposure. Models followed the same adjustment approach described above.

**Differential BP improvement.** Among survivors with uncontrolled pre-ICH BP and BP data available in both windows, we used linear regression to estimate average differences in systolic BP reduction from the pre-ICH to post-ICH window by race and deprivation, controlling for pre-ICH systolic BP. Models were additionally adjusted for age and sex, and then for neighborhood deprivation (in race models) or race/ethnicity (in deprivation models).

**Mediation analysis.** As a complementary approach to formally quantify the contribution of pre-ICH BP to post-ICH disparities, causal mediation analysis estimated the proportion of post-ICH BP disparities attributable to pre-ICH BP disparities (the indirect effect) versus factors operating after ICH (the direct effect). For each exposure (Black vs White; Deprived vs Privileged), we specified a mediator model predicting pre-ICH mean systolic BP from the exposure and covariates, and an outcome model predicting post-ICH BP from the exposure, the mediator, and covariates, using the “mediation” R package^30^ with 5,000 quasi-Bayesian simulations. We estimated the average causal mediation effect (ACME), average direct effect (ADE), total effect, and proportion mediated. We modeled both binary (controlled versus uncontrolled BP) and continuous (mean systolic BP) outcomes. Models were first adjusted for age at ICH and sex, and then additionally adjusted for neighborhood deprivation (race models) or race/ethnicity (deprivation models).

## Results

Among 2,226 ICH survivors (mean age 60.0 years; 50.6% female), 55% identified as White, 18% as Black, 16% as Hispanic/Latino, and 11% as another race or ethnicity. All participants had BP data in at least one analytic window: 79% had data before ICH (n=1,760) and 83% after ICH (n=1,852); 62% had data in both windows (n=1,386). Baseline demographic, clinical, and socioeconomic characteristics of the cohort are described stratified by pre-ICH BP status in Table 1.

**Table 1.** Baseline Cohort Characteristics Stratified by Pre-ICH Blood Pressure Status. Values under 20 were suppressed (<20) per All of Us privacy rules.

| Characteristic | Uncontrolled pre-ICH | Controlled pre-ICH | Missing pre-ICH BP | Overall |
| --- | --- | --- | --- | --- |
| **DEMOGRAPHICS** |  |  |  |  |
| **n** | 433 | 1327 | 466 | 2226 |
| Age at ICH, years, mean (SD) | 65.2 (13.1) | 59.7 (15.6) | 56.0 (15.2) | 60.0 (15.3) |
| **Sex, n (%)** |  |  |  |  |
| Male | 212 (49.0%) | 644 (48.5%) | 244 (52.4%) | 1100 (49.4%) |
| Female/Other | 221 (51.0%) | 683 (51.5%) | 222 (47.6%) | 1126 (50.6%) |
| **Race/ethnicity, n (%)** |  |  |  |  |
| White | 212 (49.0%) | 779 (58.7%) | 227 (48.7%) | 1218 (54.7%) |
| Black | 119 (27.5%) | 187 (14.1%) | 98 (21.0%) | 404 (18.1%) |
| Hispanic/Latino | 61 (14.1%) | 207 (15.6%) | 90 (19.3%) | 358 (16.1%) |
| Other | 41 (9.5%) | 154 (11.6%) | 51 (10.9%) | 246 (11.1%) |
| Missing | <20 | <20 | <20 | <20 |
| **BLOOD PRESSURE MEASUREMENTS (Pre-ICH)** |  |  |  |  |
| Mean SBP, mmHg, mean (SD) | 153.2 (13.5) | 123.9 (10.3) | – | 131.1 (16.9) |
| Number of BP measurements, median [IQR] | 7 [3–25] | 7 [3–22] | – | 7 [3–23] |
| **BLOOD PRESSURE MEASUREMENTS (Post-ICH)** |  |  |  |  |
| Mean SBP, mmHg, mean (SD) | 139.5 (14.6) | 124.9 (12.7) | 129.5 (17.2) | 128.7 (15.3) |
| Number of BP measurements, median [IQR] | 10 [5–24] | 10 [4–23] | 3 [3–11] | 8 [3–21] |
| **SOCIOECONOMIC FACTORS** |  |  |  |  |
| Deprivation index, mean (SD) [range] | 0.319 (0.064) [0.163–0.491] | 0.314 (0.071) [0.163–0.491] | 0.320 (0.063) [0.163–0.491] | 0.316 (0.068) [0.163–0.491] |
| **Deprivation tertile, n (%)** |  |  |  |  |
| Privileged | 125 (28.9%) | 448 (33.8%) | 138 (29.6%) | 711 (31.9%) |
| Intermediate | 149 (34.4%) | 424 (32.0%) | 138 (29.6%) | 711 (31.9%) |
| Deprived | 144 (33.3%) | 403 (30.4%) | 164 (35.2%) | 711 (31.9%) |
| Missing | <20 | 52 (3.9%) | 26 (5.6%) | 93 (4.2%) |
| **Annual household income, n (%)** |  |  |  |  |
| Less than \$35,000 | 165 (38.1%) | 440 (33.2%) | 161 (34.5%) | 766 (34.4%) |
| \$35,000–74,999 | 60 (13.9%) | 240 (18.1%) | 73 (15.7%) | 373 (16.8%) |
| \$75,000–99,999 | 23 (5.3%) | 92 (6.9%) | 35 (7.5%) | 150 (6.7%) |
| \$100,000 or more | 60 (13.9%) | 226 (17.0%) | 65 (13.9%) | 351 (15.8%) |
| Prefer not to answer | 125 (28.9%) | 329 (24.8%) | 132 (28.3%) | 586 (26.3%) |
| Missing | <20 | <20 | <20 | <20 |
| **Education, n (%)** |  |  |  |  |
| Did not complete high school | 71 (16.4%) | 116 (8.7%) | 62 (13.3%) | 249 (11.2%) |
| High school graduate | 103 (23.8%) | 262 (19.7%) | 114 (24.5%) | 479 (21.5%) |
| College degree or higher | 252 (58.2%) | 915 (69.0%) | 273 (58.6%) | 1440 (64.7%) |
| Prefer not to answer | <20 | 34 (2.6%) | <20 | 58 (2.6%) |
| Missing | <20 | <20 | <20 | <20 |
| **Insurance type, n (%)** |  |  |  |  |
| Medicaid | 104 (24.0%) | 292 (22.0%) | 111 (23.8%) | 507 (22.8%) |
| Medicare | 151 (34.9%) | 399 (30.1%) | 112 (24.0%) | 662 (29.7%) |
| Private/Employer | 84 (19.4%) | 372 (28.0%) | 126 (27.0%) | 582 (26.1%) |
| Other | 31 (7.2%) | 79 (6.0%) | 32 (6.9%) | 142 (6.4%) |
| Missing | 63 (14.5%) | 185 (13.9%) | 85 (18.2%) | 333 (15.0%) |
| BMI, kg/m<sup>2</sup>, mean (SD) | 30.0 (6.9) | 29.5 (7.6) | 28.3 (5.8) | 29.3 (7.1) |
| **BEHAVIORAL RISK FACTORS** |  |  |  |  |
| **Ever smoker, n (%)** |  |  |  |  |
| Yes | 200 (46.2%) | 614 (46.3%) | 223 (47.9%) | 1037 (46.6%) |
| No | 223 (51.5%) | 660 (49.7%) | 229 (49.1%) | 1112 (50.0%) |
| Prefer not to answer | <20 | 53 (4.0%) | <20 | 77 (3.5%) |
| Missing | <20 | <20 | <20 | <20 |
| **CARDIOVASCULAR RISK FACTORS** |  |  |  |  |
| Hypertension, n (%) | 358 (82.7%) | 860 (64.8%) | 183 (39.3%) | 1401 (62.9%) |
| Diabetes mellitus, n (%) | 173 (40.0%) | 385 (29.0%) | 63 (13.5%) | 621 (27.9%) |
| Hyperlipidemia, n (%) | 294 (67.9%) | 763 (57.5%) | 126 (27.0%) | 1183 (53.1%) |
| Atrial fibrillation, n (%) | 80 (18.5%) | 239 (18.0%) | 34 (7.3%) | 353 (15.9%) |
| **PRIOR CARDIOVASCULAR EVENTS** |  |  |  |  |
| Any prior cardiovascular event, n (%) | 165 (38.1%) | 364 (27.4%) | 60 (12.9%) | 589 (26.5%) |
| **OTHER COMORBIDITIES** |  |  |  |  |
| Chronic kidney disease, n (%) | 110 (25.4%) | 210 (15.8%) | 40 (8.6%) | 360 (16.2%) |
| Heart failure, n (%) | 113 (26.1%) | 273 (20.6%) | 44 (9.4%) | 430 (19.3%) |
| Dementia, n (%) | 35 (8.1%) | 80 (6.0%) | <20 | 125 (5.6%) |
| Depression, n (%) | 129 (29.8%) | 422 (31.8%) | 66 (14.2%) | 617 (27.7%) |
| COPD, n (%) | 75 (17.3%) | 212 (16.0%) | 35 (7.5%) | 322 (14.5%) |
| Cancer, n (%) | 124 (28.6%) | 415 (31.3%) | 57 (12.2%) | 596 (26.8%) |

### Pre-ICH Blood Pressure Control

In the year before ICH, 24.6% of survivors had uncontrolled BP (≥140 mmHg; N=1,760). In unadjusted analyses, uncontrolled BP was significantly more common among Black survivors (38.9%) than White survivors (21.4%; p<0.001), while rates among Hispanic/Latino (22.8%; p=0.630) and Other race and ethnicity (21.0%) survivors were similar to White survivors (Figure 2B). By neighborhood deprivation, 26.3% of deprived survivors had uncontrolled BP compared with 21.8% of privileged survivors (p=0.077), while the difference between intermediate (26.0%) and privileged survivors was also not statistically significant (p=0.097; Figure 2A).

**Figure 2.**
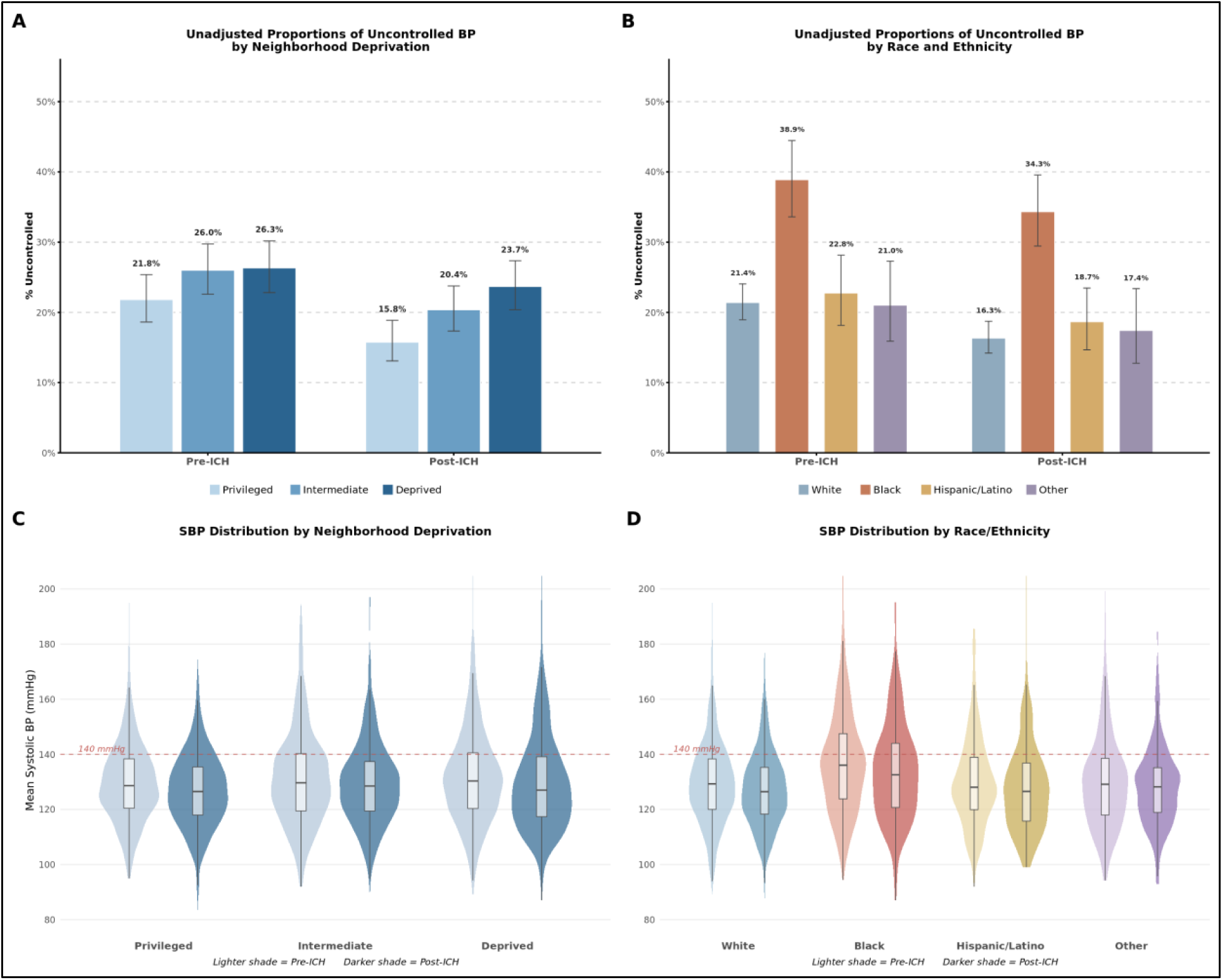
Racial and socioeconomic disparities in blood pressure control before and after intracerebral hemorrhage. (A, B) Unadjusted proportions of uncontrolled BP (systolic ≥140 mmHg) by neighborhood deprivation tertile (A) and race/ethnicity (B), with error bars indicating 95% confidence intervals. (C, D) Violin plots with embedded box plots showing distributions of mean systolic BP before (light shade) and after (dark shade) ICH, by neighborhood deprivation tertile (C) and race/ethnicity (D). The dashed line at 140 mmHg indicates the primary threshold for uncontrolled BP. BP indicates blood pressure; ICH, intracerebral hemorrhage; SBP, systolic blood pressure.

In models adjusted for age, sex, and mutually adjusted for either race/ethnicity or deprivation, Black survivors had over three times the odds of uncontrolled pre-ICH BP compared with White survivors (OR 3.16; 95% CI 2.32–4.31; P<0.001), while Hispanic/Latino (OR 1.47; 95% CI 1.03–2.09; P=0.033) survivors had modestly elevated odds and Other (OR 1.09; 95% CI 0.69–1.71; P=0.709) survivors did not differ significantly from White survivors (N=1,693; Figure 3D). Effects of intermediate (OR 1.17; 95% CI 0.88–1.57; P=0.278) and deprived (OR 1.11; 95% CI 0.82–1.51; P=0.485) neighborhoods were not statistically significant after controlling for race (N=1,693; Figure 3C).

**Figure 3.**
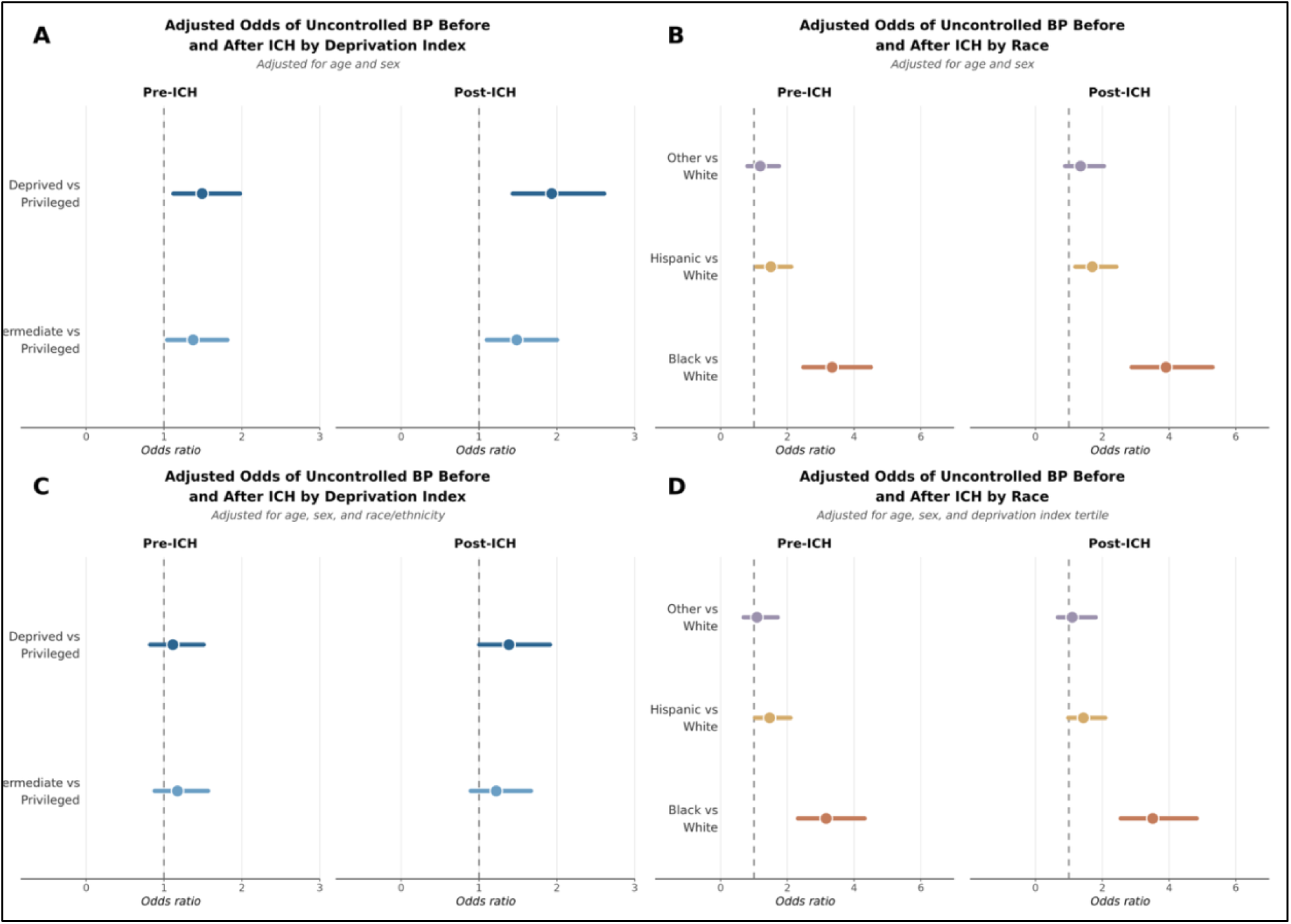
Adjusted odds of uncontrolled blood pressure before and after intracerebral hemorrhage by neighborhood deprivation and race. Forest plots showing adjusted odds ratios and 95% confidence intervals for uncontrolled BP (systolic ≥140 mmHg) before ICH (1–365 days pre-ICH) and after ICH (30–365 days post-ICH). (A) Odds of uncontrolled BP by neighborhood deprivation tertile (reference: Privileged), adjusted for age and sex. (B) Odds of uncontrolled BP by race/ethnicity (reference: White), adjusted for age and sex. (C) Odds of uncontrolled BP by neighborhood deprivation tertile, additionally adjusted for race/ethnicity. (D) Odds of uncontrolled BP by race/ethnicity, additionally adjusted for neighborhood deprivation index tertile. The vertical dashed line at 1.0 indicates no association. BP indicates blood pressure; ICH, intracerebral hemorrhage.

### Post-ICH Blood Pressure Control

Between 30 and 365 days after ICH, 20.1% of survivors had uncontrolled BP (N=1,852). Uncontrolled BP was significantly more common among Black survivors (34.3%) than White survivors (16.3%; p<0.001), while rates among Hispanic/Latino (18.7%; p=0.344) and Other race/ethnicity (17.4%) survivors did not differ significantly from White survivors (Figure 2B). By neighborhood deprivation, both intermediate (20.4%; p=0.037) and deprived (23.7%; p<0.001) survivors had significantly higher rates of uncontrolled BP compared with privileged survivors (15.8%; Figure 2A).

In models adjusted for age, sex, and mutually adjusted for either race/ethnicity or deprivation, Black ICH survivors were more than three times more likely than White survivors to have uncontrolled BP (OR 3.51; 95% CI 2.55–4.83; P<0.001). Hispanic/Latino (OR 1.43; 95% CI 0.98–2.09; P=0.061) and Other (OR 1.10; 95% CI 0.67–1.80; P=0.708) ICH survivors did not have significantly greater odds of uncontrolled BP than White survivors (N=1,783; Figure 3D). Survivors from deprived neighborhoods had significantly greater odds of uncontrolled BP after ICH than privileged survivors (OR 1.38; 95% CI 1.00–1.91; P=0.048), while the intermediate comparison was not statistically significant (OR 1.22; 95% CI 0.89–1.67; P=0.208; N=1,783; Figure 3C). In a joint exposure analysis, Black survivors from deprived neighborhoods had over four times the odds of uncontrolled post-ICH BP compared with White survivors from privileged neighborhoods (OR 4.24; 95% CI 2.73–6.59; P<0.001; N=912).

### Sensitivity Analyses

Racial disparities in BP control persisted across all sensitivity thresholds (120, 130, 140, and 150 mmHg). In models adjusted for age, sex, and neighborhood deprivation, odds of uncontrolled BP for Black versus White survivors ranged from OR 1.93 to 3.44 before ICH and from OR 2.17 to 3.51 after ICH across these thresholds (all P<0.001). Hispanic/Latino survivors had significantly elevated odds of uncontrolled pre-ICH BP at 140 mmHg (OR 1.47; P=0.033) and 150 mmHg (OR 1.62; P=0.043), and at 150 mmHg post-ICH (OR 1.84; P=0.016). In models adjusted for age, sex, and race/ethnicity, deprived neighborhoods were associated with significantly higher odds of uncontrolled post-ICH BP at 140 and 150 mmHg (OR 1.38, P=0.048 and OR 2.32, P<0.001, respectively), but no pre-ICH disparities were significant when adjusting for race/ethnicity.

In a sensitivity analysis restricting the cohort to intraparenchymal hemorrhage only (N=1,108), racial disparities in BP control were consistent with the primary analysis (Black vs White: OR 3.00, P<0.001 pre-ICH; OR 3.48, P<0.001 post-ICH), and pre-ICH BP mediated 23% of the Black-White disparity in uncontrolled post-ICH BP (P<0.001). Neighborhood deprivation associations were directionally consistent but did not reach statistical significance in fully adjusted models.

### Change in Blood Pressure Control Surrounding ICH

Among the 1,386 survivors with BP data in both analytic windows, 331 (23.9%) had uncontrolled pre-ICH BP and 1,055 (76.1%) had controlled pre-ICH BP. Overall, 56.8% of previously uncontrolled survivors achieved control after ICH, and 11.3% of previously controlled survivors lost control.

***Achievement of BP Control***. Among survivors with uncontrolled pre-ICH BP, 66.7% of White survivors achieved BP control after ICH compared with 44.8% of Black survivors (p=0.001), 52.4% of Hispanic/Latino survivors (p=0.085), and 46.4% of survivors of other race and ethnicities (N=331; Figure 4A). In models adjusted for age and sex, Black survivors had significantly lower odds of achieving control (OR 0.37; 95% CI 0.21–0.65; P<0.001; N=331; Figure 4C). By neighborhood deprivation, 69.1% of privileged, 51.7% of intermediate (p=0.010), and 52.8% of deprived (p=0.018) survivors achieved control (N=319; Figure 4B). In models adjusted for age and sex, both intermediate (OR 0.48; 95% CI 0.27–0.86; P=0.013) and deprived (OR 0.50; 95% CI 0.28–0.89; P=0.020) survivors had significantly lower odds of achieving control (Figure 4D), though these associations were attenuated to nonsignificant after additional adjustment for race (N=319).

**Figure 4.**
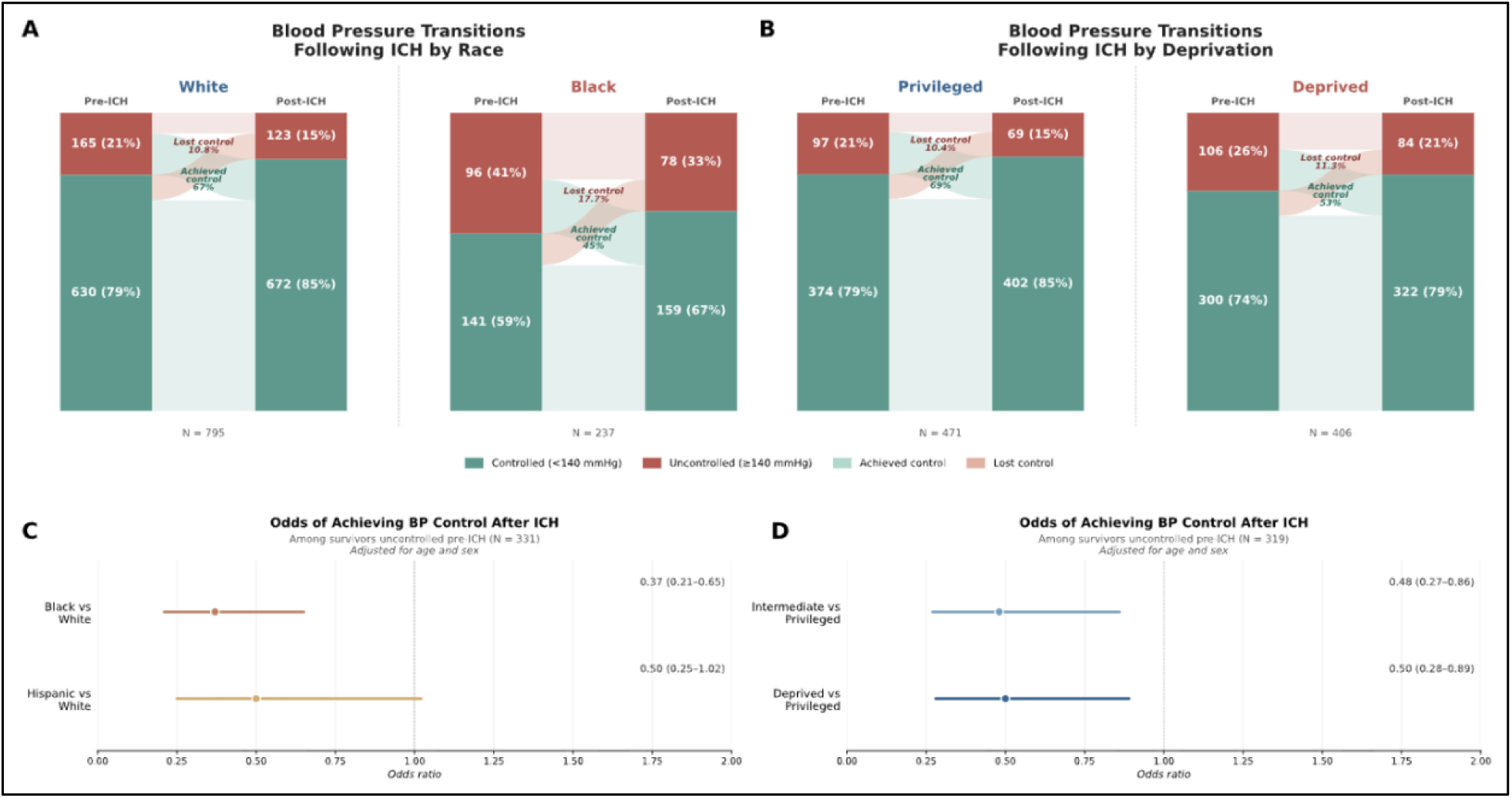
Blood pressure transitions following intracerebral hemorrhage and odds of achieving control by race and deprivation. (A, B) Stacked bar plots showing blood pressure transitions from pre-ICH to post-ICH among survivors with data in both analytic windows, by race (A; White vs Black) and neighborhood deprivation (B; Privileged vs Deprived). Each pair of bars shows the proportion with controlled (<140 mmHg, green) and uncontrolled (≥140 mmHg, red) BP before and after ICH, with lighter shades indicating transitions: achieved control (light green) and lost control (light red). (C, D) Forest plots showing adjusted odds ratios and 95% confidence intervals for achieving BP control after ICH among survivors with uncontrolled pre-ICH BP (N=319), by race (C; reference: White; adjusted for age and sex) and neighborhood deprivation (D; reference: Privileged; adjusted for age and sex). Odds ratios below 1.0 indicate lower likelihood of achieving control. BP indicates blood pressure; ICH, intracerebral hemorrhage.

***Loss of BP Control.*** Among survivors with controlled pre-ICH BP, 10.8% of White survivors lost control compared with 17.7% of Black survivors (p=0.022), 9.5% of Hispanic/Latino survivors (p=0.634), and 8.6% of survivors of other race and ethnicities (N=1,055). In fully adjusted models Black survivors had nearly three times the odds of losing control (OR 2.76; 95% CI 1.57–4.85; P<0.001; N=1,024). By neighborhood deprivation, intermediate (OR 1.15; 95% CI 0.70–1.88; P=0.576) and deprived (OR 1.12; 95% CI 0.65–1.93; P=0.673) neighborhoods were not significantly associated with loss of control (N=1,024).

### Differential Blood Pressure Improvement After ICH

Among the 331 survivors with uncontrolled pre-ICH BP, White survivors had a mean systolic BP reduction of 14.2 mmHg (95% CI 12.3–16.1) compared with 11.5 mmHg (95% CI 8.0–15.0) for Black, 7.3 mmHg (95% CI 2.0–12.5) for Hispanic/Latino, and 13.3 mmHg (95% CI 7.9–18.7) for Other survivors. By neighborhood deprivation, privileged survivors had a mean reduction of 14.1 mmHg (95% CI 11.4–16.9) compared with 11.6 mmHg (95% CI 8.5–14.8) for intermediate and 12.3 mmHg (95% CI 9.6–15.0) for deprived survivors.

In models adjusted for pre-ICH systolic BP, age, and sex, Black survivors achieved 7.7 mmHg less systolic BP improvement than White survivors starting at the same pre-ICH BP (95% CI 3.9–11.5; P<0.001), and Hispanic/Latino survivors achieved 8.8 mmHg less improvement (95% CI 4.1–13.4; P<0.001; N=331; Figure 5A). Intermediate (5.8 mmHg; 95% CI 2.0–9.7; P=0.003) and deprived (4.5 mmHg; 95% CI 0.7–8.3; P=0.022) survivors also achieved significantly less BP improvement than privileged survivors after controlling for pre-ICH systolic BP, age, and sex (N=319; Figure 5B). In models additionally adjusting for both race/ethnicity and deprivation simultaneously, racial disparities remained significant while deprivation effects became nonsignificant. Results were similar in sensitivity analyses including all survivors with pre-ICH systolic BP ≥130 mmHg, with Black survivors achieving 4.9 mmHg less improvement than White survivors (95% CI 2.4–7.5; P<0.001; N=692; Figure 5C), while the Deprived-Privileged difference was not statistically significant at this threshold (N=667; Figure 5D).

**Figure 5.**
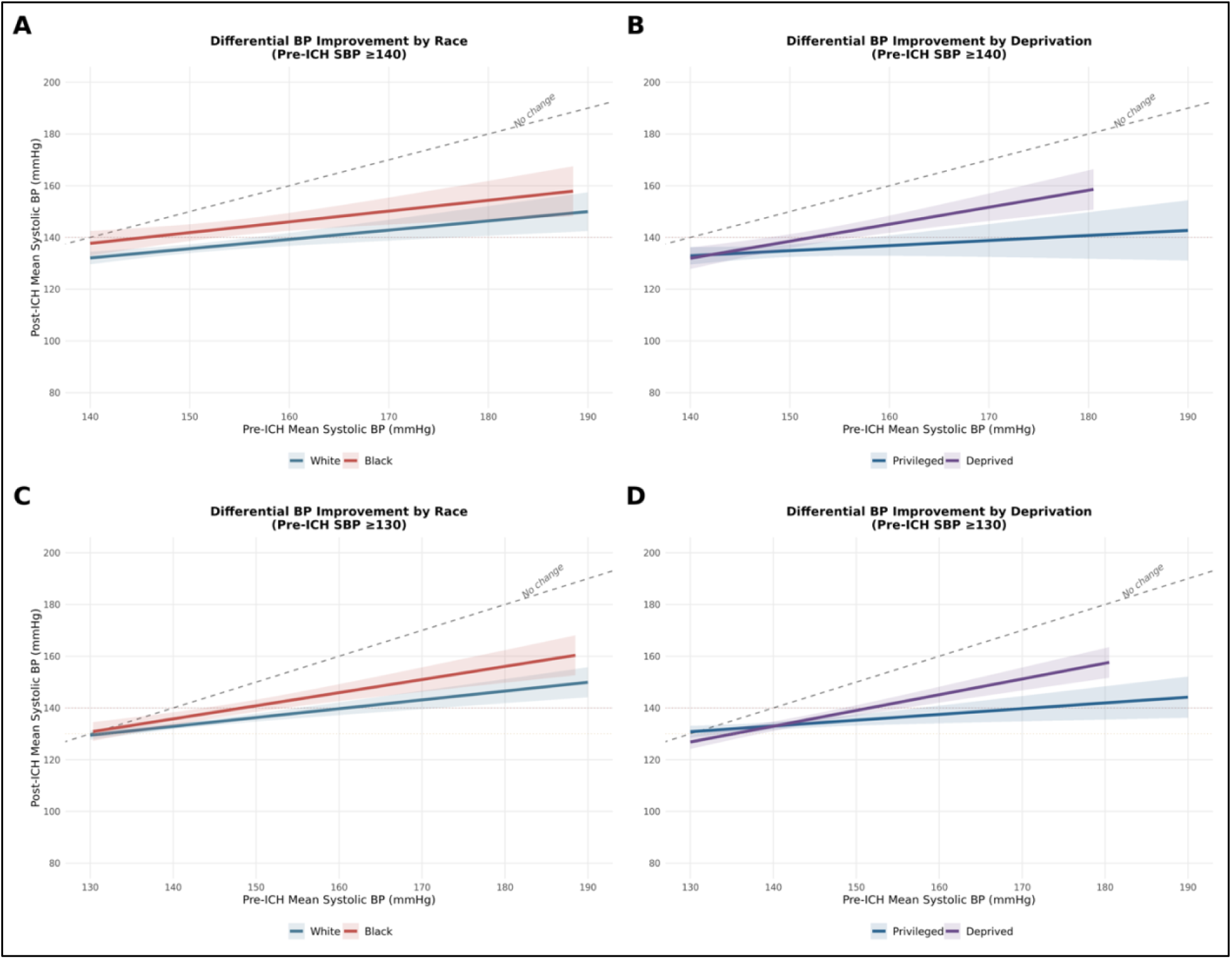
Differential blood pressure improvement after intracerebral hemorrhage by race and neighborhood deprivation. Scatter plots with linear regression lines and 95% confidence bands showing the relationship between pre-ICH and post-ICH mean systolic BP among survivors with uncontrolled pre-ICH BP. The diagonal dashed line represents no change from pre- to post-ICH; points and regression lines below this line indicate BP improvement. (A, B) Survivors with pre-ICH mean systolic BP ≥140 mmHg, by race (A; White vs Black) and neighborhood deprivation (B; Privileged vs Deprived). (C, D) Survivors with pre-ICH mean systolic BP ≥130 mmHg, by race (C) and neighborhood deprivation (D). The horizontal dotted line at 140 mmHg indicates the point at which there would be no reduction in BP after ICH. BP indicates blood pressure; ICH, intracerebral hemorrhage; SBP, systolic blood pressure.

### Contribution of Pre-ICH Blood Pressure to Post-ICH Disparities

To assess whether post-ICH disparities are explained by pre-existing BP differences, we additionally adjusted post-ICH models for pre-ICH BP control status among the 1,386 survivors with data in both windows. In models adjusted for age and sex, despite pre-ICH uncontrolled BP being a strong predictor of post-ICH uncontrolled BP (OR 4.70; 95% CI 3.46–6.38; P<0.001), adjusting for pre-ICH BP control status only modestly attenuated the Black-White disparity in uncontrolled post-ICH BP, from OR 4.05 (95% CI 2.81–5.82) to OR 2.95 (95% CI 2.00–4.35; P<0.001), indicating that the majority of the racial disparity in post-ICH BP control is not explained by pre-existing BP differences. For deprivation, the deprived-privileged disparity was attenuated from OR 1.84 (95% CI 1.28–2.63) to OR 1.64 (95% CI 1.12–2.40; P=0.011) after adjusting for pre-ICH BP status, age, and sex. After additionally adjusting for race, the deprivation association was attenuated to nonsignificant (OR 1.26; 95% CI 0.84–1.89; P=0.271), suggesting that neighborhood deprivation disparities in post-ICH BP are largely explained by pre-existing BP differences and their correlation with race.

To formally quantify the contribution of pre-ICH BP to post-ICH disparities, we conducted causal mediation analyses. In models adjusted for age and sex, pre-ICH BP mediated 27% (95% CI 17–39%; P<0.001) of the Black-White disparity in uncontrolled post-ICH BP (ACME 0.07; 95% CI 0.04–0.10; N=1,032; Figure 6D) and 57% of the racial disparity in mean post-ICH systolic BP (ACME 5.00 mmHg; 95% CI 3.78–6.25; N=1,032; Figure 6B). Pre-ICH BP mediated 26% (95% CI 6–58%; P=0.014) of the deprived-privileged disparity in uncontrolled post-ICH BP (ACME 0.02; 95% CI 0.01–0.04; N=877; Figure 6C) and 65% of the disparity in mean post-ICH systolic BP (ACME 1.90 mmHg; 95% CI 0.85–2.96; N=877; Figure 6A). After mutual adjustment for the other exposure, race mediation results were unchanged (27% mediated; P<0.001; N=1,031), while the deprivation ACME was attenuated to nonsignificant (P=0.148; N=877).

**Figure 6.**
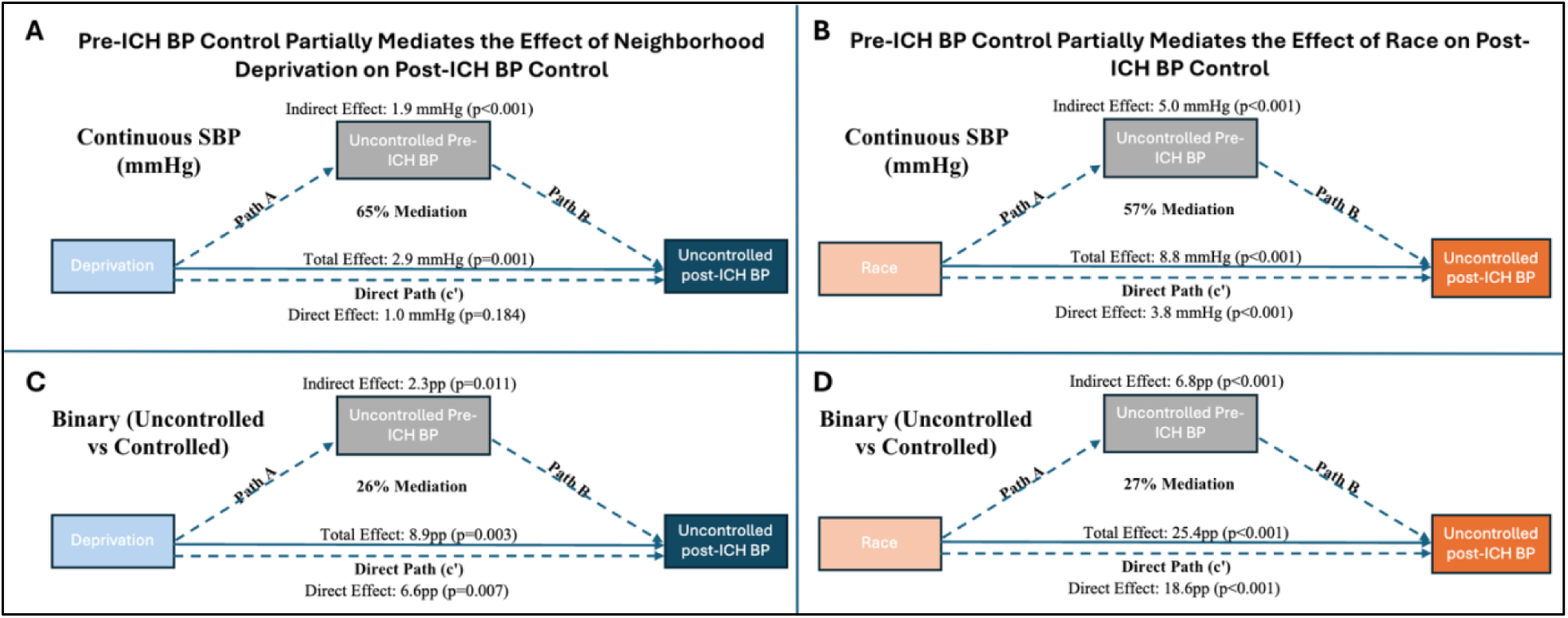
Pre-ICH blood pressure mediates disparities in post-ICH control. Mediation analysis results showing the proportion of racial and socioeconomic disparities in post-ICH BP outcomes attributable to pre-ICH BP control. (A, B) Mediation of disparities in continuous mean post-ICH systolic BP by neighborhood deprivation (A) and race (B). (C, D) Mediation of disparities in binary uncontrolled post-ICH BP (systolic ≥140 mmHg) by neighborhood deprivation (C) and race (D). Solid arrows represent direct effects; dashed arrows represent indirect effects mediated by pre-ICH BP. ACME indicates average causal mediation effect; BP, blood pressure; DI, Deprivation Index; pp, percentage points; SBP, systolic blood pressure. All models adjusted for age and sex.

## Discussion

In this national cohort of ICH survivors, we found substantial racial and socioeconomic disparities in blood pressure control before and after ICH. Black survivors had over threefold odds of uncontrolled post-ICH BP compared with White survivors, a disparity that was consistent across thresholds from 120 to 150 mmHg and persisted after adjustment for neighborhood deprivation. ICH survivors living in deprived neighborhoods also had significantly higher odds of uncontrolled post-ICH BP. Despite pre-ICH uncontrolled BP being a strong predictor of post-ICH uncontrolled BP, adjusting for pre-ICH BP control status only modestly attenuated both the racial and deprivation disparities, and mediation analyses confirmed that pre-ICH BP explained only 27% of the racial disparity and 26% of the deprivation disparity. Change-in-status analyses showed robust racial differences in post-ICH BP trajectories: Black survivors were less likely to achieve control and more likely to lose control after ICH. Socioeconomic differences followed a similar pattern in unadjusted and partially adjusted analyses, though several deprivation effects were attenuated after adjustment for race/ethnicity. These findings suggest that the post-ICH period represents a window in which disparities in BP management actively expand pre-existing inequities.

Our results extend prior work documenting racial and socioeconomic disparities in hypertension management after stroke. In a single-center cohort of 336 ICH patients, Abramson et al. found that racial and ethnic minority ICH survivors were significantly less likely to experience BP reductions in the year following ICH than White survivors. However, pre-stroke BP control status predicted post-stroke BP status regardless of race and socioeconomic status, raising the question of whether post-ICH disparities primarily reflect pre-existing inequities or differential post-stroke management. Our study addressed that question by directly adjusting for pre-ICH BP control status in a larger, more diverse cohort, with mediation analysis as a complementary approach. The answer appears to be that both pre-existing inequalities and post-stroke factors play a significant role, but differences in post-stroke BP management drive most of the disparity.

The change-in-status and differential BP improvement analyses provide additional evidence for this interpretation. Among survivors who were uncontrolled before ICH, 67% of White survivors achieved BP control afterward compared with only 45% of Black survivors, and White survivors experienced approximately 8 mmHg more systolic BP reduction than Black survivors after adjustment for baseline BP, age, and sex. Among survivors who were controlled before ICH, Black survivors were also nearly three times as likely to lose control. Neighborhood deprivation showed a similar pattern with privileged survivors being significantly more likely to achieve control and experiencing significantly greater BP reductions than intermediate and deprived survivors. These results indicate that the post-ICH period differentially benefits more advantaged populations, who appear to receive more effective hypertension management, while disadvantaged populations are less likely to improve and more likely to deteriorate.

Our observation that a substantial portion of post-ICH BP disparities reflects pre-existing inequities underscores the need for population-level interventions to improve hypertension control in Black adults and residents of socioeconomically disadvantaged communities. Without addressing the structural barriers driving these inequities, fully eliminating post-ICH disparities may not be possible. However, the finding that most of the post-ICH disparity in rates of BP control is not explained by pre-existing differences suggests that much of the burden of recurrent ICH, disability, and mortality attributable to uncontrolled post-ICH BP among disadvantaged populations may be addressable through targeted improvements in post-stroke care. The fact that more advantaged populations consistently achieve BP control after ICH at substantially higher rates demonstrates that effective post-ICH BP management is achievable. The challenge is to understand why current approaches are less effective for Black and socioeconomically deprived survivors and to develop interventions that reduce the gap.

Several lines of evidence suggest that this is a modifiable problem. A standardized post-stroke management protocol reduced some racial differences in vascular risk factor management in a controlled clinical setting,^19,20^ and emerging interventions including telehealth-based home BP monitoring have shown early promise in a pilot trial among underserved stroke survivors,^31^ though evidence from larger trials of coordinated care models has been mixed.^32^ Our findings provide population-level evidence that much of the post-ICH BP disparity facing disadvantaged groups emerges during the post-stroke treatment period rather than being carried over from pre-existing differences. These results suggest that such interventions could substantially reduce disparities at a broader scale, though achieving that potential requires better understanding of the specific treatment differences affecting disadvantaged populations after ICH. Future research should aim to better characterize disparities in antihypertensive prescribing patterns, medication adherence, access to follow-up, and other structural factors that may lead to worse BP outcomes for minority and socioeconomically deprived populations. Investing in post-stroke care disparity research and interventions that standardize BP management after ICH could meaningfully close the gap in post-ICH outcomes.

This study has several strengths, including the use of a large, racially and socioeconomically diverse national cohort with longitudinal EHR-derived BP data spanning both the pre-ICH and post-ICH periods within the same individuals, multiple complementary analytic approaches, and analyses across multiple systolic BP thresholds. This study also has important limitations. The observational design prevents causal inference about specific mechanisms driving post-ICH BP disparities. We did not include data about ICH severity measures which may confound results if severity differs by race or socioeconomic status. We also did not analyze antihypertensive medication data or other factors related to BP management, preventing direct examination of treatment differences. Neighborhood deprivation was linked via 3-digit ZIP codes, which represent broad geographic areas and may not capture the granularity of neighborhood-level socioeconomic context. The inclusion of only ICH patients surviving at least one year with available BP data introduces survivor and availability biases, potentially excluding the most severely affected patients and those with the least access to healthcare. Finally, the Other race and ethnicity category combines heterogeneous groups due to sample size constraints, limiting our ability to characterize disparities within certain racial and ethnic populations.

## Conclusions

In a national cohort, racial and socioeconomic disparities in BP control are present before ICH and persist afterward, but the majority of post-ICH disparities do not seem to be explained by pre-existing differences. Instead, more advantaged populations consistently achieve greater BP improvement after ICH, while disadvantaged populations are less likely to improve during the post-ICH management period. Because effective post-ICH BP management is demonstrably achievable in more advantaged populations, targeted interventions that identify and address the specific barriers to BP control in Black and socioeconomically deprived ICH survivors have the potential to substantially reduce the burden of recurrent hemorrhage and close persistent BP disparities in these disadvantaged populations.

## Data Availability

Data used in this study are available from the All of Us Research Program (https://allofus.nih.gov/) for researchers with controlled tier access.

## Acknowledgements

We gratefully acknowledge All of Us participants for their contributions, without whom this research would not have been possible. We also thank the National Institutes of Health’s All of Us Research Program for making available the participant data examined in this study.

## Non-standard Abbreviations and Acronyms

ACME: Average Causal Mediation Effect
ADE: Average Direct Effect
BP: Blood Pressure
SBP: Systolic Blood Pressure
CI: Confidence Interval
EHR: Electronic Health Records
ICH: Intracerebral Hemorrhage
OR: Odds Ratio

## References

1. PROGRESS Collaborative Group. Randomised trial of a perindopril-based blood-pressure-lowering regimen among 6105 individuals with previous stroke or transient ischaemic attack. Lancet. 2001;358(9287):1033–1041.

2. SPS3 Study Group, Benavente OR, Coffey CS, et al. Blood-pressure targets in patients with recent lacunar stroke: the SPS3 randomised trial. Lancet. 2013;382(9891):507–515.

3. Biffi A, Anderson CD, Battey TWK, et al. Association between blood pressure control and risk of recurrent intracerebral hemorrhage. JAMA. 2015;314(9):904–912.

4. Zahuranec DB, Wing JJ, Edwards DF, et al. Poor long-term blood pressure control after intracerebral hemorrhage. Stroke. 2012;43(10):2580–2585.

5. Kalasapudi L, Williamson S, Shipper AG, et al. Scoping review of racial, ethnic, and sex disparities in the diagnosis and management of hemorrhagic stroke. Neurology. 2023;101(3):e267–e276.

6. Carey RM, Whelton PK. Prevention, detection, evaluation, and management of high blood pressure in adults: synopsis of the 2017 American College of Cardiology/American Heart Association Hypertension Guideline. Ann Intern Med. 2018;168(5):351–358.

7. Carnethon MR, Pu J, Howard G, et al. Cardiovascular health in African Americans: a scientific statement from the American Heart Association. Circulation. 2017;136(21):e393–e423.

8. Wyatt SB, Akylbekova EL, Wofford MR, et al. Prevalence, awareness, treatment, and control of hypertension in the Jackson Heart Study. Hypertension. 2008;51(3):650–656.

9. Hertz RP, Unger AN, Cornell JA, Saunders E. Racial disparities in hypertension prevalence, awareness, and management. Arch Intern Med. 2005;165(18):2098–2104.

10. Aggarwal R, Chiu N, Wadhera RK, et al. Racial/ethnic disparities in hypertension prevalence, awareness, treatment, and control in the United States, 2013 to 2018. Hypertension. 2021;78(6):1719–1726.

11. Muntner P, Hardy ST, Fine LJ, et al. Trends in blood pressure control among US adults with hypertension, 1999-2000 to 2017-2018. JAMA. 2020;324(12):1190–1200.

12. Claudel SE, Adu-Brimpong J, Banks A, et al. Association between neighborhood-level socioeconomic deprivation and incident hypertension: a longitudinal analysis of data from the Dallas Heart Study. Am Heart J. 2018;204:109–118.

13. Saban MT, Tootooni S, Markossian TW, et al. The association of area deprivation index and blood pressure control and therapeutic inertia among older adults with hypertension. J Hum Hypertens. 2025;39(11):748–754.

14. Akinyelure OP, Jaeger BC, Oparil S, et al. Social determinants of health and uncontrolled blood pressure in a national cohort of Black and White US adults: the REGARDS Study. Hypertension. 2023;80(7):1403–1413.

15. Akinyelure OP, Jaeger BC, Moore TL, et al. Racial differences in blood pressure control following stroke: the REGARDS Study. Stroke. 2021;52(12):3944–3952.

16. Cruz-Flores S, Rabinstein A, Biller J, et al. Racial-ethnic disparities in stroke care: the American experience: a statement for healthcare professionals from the American Heart Association/American Stroke Association. Stroke. 2011;42(7):2091–2116.

17. Levine DA, Neidecker MV, Kiefe CI, Karve S, Williams LS, Allison JJ. Racial/ethnic disparities in access to physician care and medications among US stroke survivors. Neurology. 2011;76(1):53–61.

18. Stamm B, Royan R, Cui J, et al. Trends in Black-White differences of antihypertensive treatment in individuals with and without history of stroke. Stroke. 2024;55(8):2034–2044.

19. Almallouhi E, Nelson AM, Cotsonis G, Harris W, Chimowitz MI, Turan TN. Ameliorating racial disparities in vascular risk factor management with aggressive medical management in the SAMMPRIS trial. Stroke. 2023;54(9):2235–2240.

20. Chimowitz MI, Lynn MJ, Derdeyn CP, et al. Stenting versus aggressive medical therapy for intracranial arterial stenosis. N Engl J Med. 2011;365(11):993–1003.

21. Woo D, Rosand J, Kidwell C, et al. The Ethnic/Racial Variations of Intracerebral Hemorrhage (ERICH) Study Protocol. Stroke. 2013;44(10):e120–e125.

22. Rodriguez-Torres A, Murphy M, Kourkoulis C, et al. Hypertension and intracerebral hemorrhage recurrence among White, Black, and Hispanic individuals. Neurology. 2018;91(1):e37–e44.

23. Leasure AC, King ZA, Torres-Lopez V, et al. Racial/ethnic disparities in the risk of intracerebral hemorrhage recurrence. Neurology. 2020;94(3):e314–e322.

24. Abramson JR, Castello JP, Keins S, et al. Biological and social determinants of hypertension severity before vs after intracerebral hemorrhage. Neurology. 2022;98(13):e1349–e1360.

25. All of Us Research Program Investigators, Denny JC, Rutter JL, et al. The “All of Us” Research Program. N Engl J Med. 2019;381(7):668–676.

26. Brokamp C, Beck AF, Goyal NK, Ryan P, Greenberg JM, Hall ES. Material community deprivation and hospital utilization during the first year of life: an urban population-based cohort study. Ann Epidemiol. 2019;30:37–43.

27. Sakhuja S, Colvin CL, Akinyelure OP, et al. Reasons for uncontrolled blood pressure among US adults: data from the US National Health and Nutrition Examination Survey. Hypertension. 2021;78(5):1567–1576.

28. Chobanian AV, Bakris GL, Black HR, et al. Seventh report of the Joint National Committee on Prevention, Detection, Evaluation, and Treatment of High Blood Pressure. Hypertension. 2003;42(6):1206–1252.

29. R Core Team. R: A Language and Environment for Statistical Computing. Vienna, Austria: R Foundation for Statistical Computing; 2024.

30. Tingley D, Yamamoto T, Hirose K, Keele L, Imai K. mediation: R package for causal mediation analysis. J Stat Softw. 2014;59(5):1–38.

31. Naqvi IA, Strobino K, Cheung YK, et al. Telehealth After Stroke Care pilot randomized trial of home blood pressure telemonitoring in an underserved setting. Stroke. 2022;53(12):3538–3547.

32. Towfighi A, Cheng EM, Ayala-Rivera M, et al. Effect of a coordinated community and chronic care model team intervention vs usual care on systolic blood pressure in patients with stroke or transient ischemic attack: the SUCCEED randomized clinical trial. JAMA Netw Open. 2021;4(2):e2036227.

